# Can age-distribution be an indicator of the goodness of COVID-19 testing?

**DOI:** 10.1101/2020.12.21.20248690

**Authors:** Amirhoshang Hoseinpour Dehkordi, Reza Nemati, Pouya Tavousi

## Abstract

It has been evident that the faster, more accurate, and more comprehensive testing can help policymakers assess the real impact of COVID-19 and help them with when and how strict the mitigation policies should be. Nevertheless, the exact number of infected ones could not be measured due to the lack of comprehensive testing. In this paper, first of all, we will investigate the relation of transmission of COVID-19 with age by observing timed data in multiple countries. Then, we compare the COVID-19 CFR with the age-demography data. and as a result, we have proposed a method for estimating a lower bound for the number of positive cases by using the reported data on the oldest age group and the regions’ population age-distributions. The proposed estimation method improved the expected similarity between the age-distribution of positive cases and regions’ populations. Thus, using the publicly accessible data for several developed countries, we show how the improvement of testing over the course of several months has made it clear for the community that different age groups are equally prone to becoming COVID positive. The result shows that the age demography of COVID-19 gets similar to the age-demography of the population, together with the reduction of CFR over time. In addition, countries with less CFR have more similar COVID-19’s age-distribution, which is caused by more comprehensive testing, than ones who have higher CFR. This leads us to a better estimation for positive cases in different testing strategies. Having knowledge of this fact helps policymakers enforce more effective policies for controlling the spread of the virus.

## 1 Introduction

Correct evaluation of the number of confirmed cases during a pandemic is so important in order to make non-pharmaceutical interventions (NPIs). Many policies such as restricting population, travel restriction, and closing schools had benefits of controlling the transmission of the COVID-19 [1–3], but they also had real impacts on social activities. The absence of policies had also caused “exponential growth rates of approximately 38% per day” [4]. Although the policies would have different impacts on different countries in many cases, this shows the importance of NPIs. The most influential key factor in making policies could be the number of infected cases in that region. So the more accuracy in such evaluations could be mandatory. But the evaluation of such quantities always had errors. Many reasons could cause these errors, such as no symptoms transmitters, limitation in tests and etc.

The performance of any quantitative analysis focused on CFR evaluations is also influenced by our understanding of the mechanisms, by which the virus spreads. Ongoing research by the community has shed light on the reality of such mechanisms. For example, it is now evident that respiratory, contact, and aerosol transmission are the main mechanisms by which the virus spreads [5, 6]. Accordingly, countries that were heavily impacted by COVID-19 were forced to put in place various mitigation policies. Also, it has been shown that initial viral load is important for more effective transmission of the disease [5, 7]. However, there is little known about whether people of different ages have different susceptibility to the infection [8]. Given a higher mortality rate for older cases, in one study, Li et al. [5] showed that more than 50% of early Patients with COVID-19 in Wuhan were more than 60 years old. However, the under-representation of younger people was suspected to be attributed to the fact that some of the young infected people were asymptomatic [6, 9, 10]. Therefore, in the absence of a true picture of the disease behavior, data scientists are instead urged to practice more realistic assumptions when evaluating positive cases and following that the CFR.

Herein, first, we compared the relationship between some countries’ population demography with COVID-19’s population demography in the respective country, over time. This shows us how this relationship changes over time. Then, we attempt to show how a demographical comparison of COVID-19 confirmed cases and populations of different regions/countries can shed light on estimated CFR discrepancies for different regions. Also, to improve the estimation of the number of positive cases, the data on the oldest age group, namely >80-year-olds, are used to calculate an infected population ratio. This infected population ratio will then be used to estimate the number of positive cases in other age groups. In accomplishing so, the following assumptions are made: (1) In estimating the number of positive cases, the level of infection is not taken into consideration; (2) As a result of section 3.1, susceptibility to catching the virus is not age-dependent; (3) On average, >80-year-old cases have less number of social contacts than younger population [11]; and (4) >80-year-old cases are more symptomatic and with a higher probability correctly identified. Because of our last two assumptions, our analysis provides only lower bound estimates to the positive number of cases.

## 2 Materials and Methods

The data for the number of confirmed cases or deaths were taken from [12]. Also, the age-stratified data for confirmed cases were obtained from [13–18], and geographical regions’ populations’ data, available in [19]. To improve the estimation of the total number of COVID-19 confirmed cases, the following assumptions were made: (1) In estimating the number of positive cases, the level of infection is not taken into consideration; (2) Susceptibility to infection, due to contact with a positive case, is independent of age (it could be derived from 3.1); and (3) On average, >80-year-old cases have less number of social contacts than younger population [11]; (4) Data on the >80 age group more accurately reflects the demography of COVID-19 because the fraction of symptomatic cases overall positive cases is highest in this group; Using the data for >80-year-old cases, more accurate lower-bound estimates of positive cases were made for different regions, namely Spain, Italy, Germany, and South Korea. The age-distributions of estimated positive cases and confirmed cases, in regions with high and low CFRs, were compared against each other and the population’s age-distribution, for South Korea, Germany, Italy, and Spain.

In this study, first, we normalized the age-distribution of COVID-19 and population (divided by the sum) and then we use *L*_1_ norm to calculate the Differences between COVID-19’s and Population’s age-distribution (DCP). Let vector *P* = (*p*_0_, …, *p*_*n*_) as population age-distribution, and *C* = (*c*_0_, …, *c*_*n*_) as COVID-19 age-distribution. The *L*_1_ distance calculated as follows:

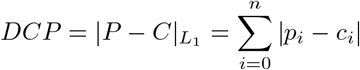

## 3 Results and Discussion

In this article, first, we will demonstrate the relation of COVID-19’s age-distribution with respective countries’ age-distribution of populations over time. Further, this work addresses the inconsistencies observed in COVID-19 CFR reported in different geographical regions.

### 3.1 Timed reports

In this section, we are going to compare the DCP of three hotspot countries, the US, Italy, and England, over time. The value of DCP here represents the difference of all population age-distribution with COVID-19’s age-distribution. As it could be observed, in all these countries the value of DCP reduces over time in all of these countries. The daily tests per thousand people in the US increases from <0.01 on March 8 to 2.54 on August 31, this shows that the more comprehensive testing results in more similarities between population age-distribution and COVID-19 age-distribution. In fig. 1 the age-distribution of COVID-19 and population and its similarity could be observed, the calculated values for the US also could be observed in table 1. The same trend for the value of DCP could be observed for Italy. The last column of Table 2 shows the daily COVID-19 cases of Italy in the collection date. These values together with the drawn plot in Figure 4b, shows that less symptomatic people tested more during the time. The figure 4b shows the trend of changing COVID-19’s age-distribution. As it could be observed, and calculated, the COVID-19’s age-distribution in Figure 2h is obviously more similar to population’s age-distribution than 2a, and the trend is monotonic. Similar analysis could be done for weekly reports of England for more exact analysis. By calculation of the DCP per week of England, it could be observed that the trend of the value of DCP is descending over time (see table 3). The plot for comparing COVID-19’s age-distribution and population’s age-distribution is depicted in Figure 3. The data provided by England’s national health institution is in the smaller time-frame, so the similarity of COVID-19 and population of age over time could be observed more easily.

**Table 1.** Calculated value of DCP for 4 different months and the relation of this value with testing in the US.

| Country | Collection Date | DCP | Test Per 1000 |
| --- | --- | --- | --- |
| The US | May | 0.32 | 0.79 |
| The US | June | 0.23 | 1.44 |
| The US | July | 0.16 | 2.26 |
| The US | Aug | 0.14 | 2.54 |

**Table 2.** Calculated value of DCP and confirmed cases for 8 different months in Italy.

| Country | Collection Date | DCP | Daily cases |
| --- | --- | --- | --- |
| Italy | March 31 | 0.58 | 4053 |
| Italy | 26 Apr | 0.53 | 2325 |
| Italy | 26 May | 0.51 | 397 |
| Italy | 30 June | 0.499 | 142 |
| Italy | 28 July | 0.483 | 181 |
| Italy | 25 Aug | 0.436 | 876 |
| Italy | 22 Sep | 0.34 | 1392 |
| Italy | 13 Oct | 0.27 | 5901 |

**Table 3.** Calculated value of DCP and confirmed cases for 27 different months in England.

| Country | Collection Date | Confirmed Cases (CC) | DCP |
| --- | --- | --- | --- |
| England | Week 13<sup>+</sup> | 25133 | 0.7212 |
| England | Week 14 | 50912 | 0.6918 |
| England | Week 15 | 77422 | 0.6299 |
| England | Week 16 | 104265 | 0.5805 |
| England | Week 17 | 131586 | 0.5471 |
| England | Week 18 | 157540 | 0.5248 |
| England | Week 19 | 176317 | 0.5106 |
| England | Week 20 | 192790 | 0.4966 |
| England | Week 21 | 206427 | 0.4874 |
| England | Week 22 | 215763 | 0.4815 |
| England | Week 23 | 223165 | 0.4762 |
| England | Week 24 | 229637 | 0.4688 |
| England | Week 25 | 235373 | 0.4617 |
| England | Week 26 | 239763 | 0.4552 |
| England | Week 27 | 243539 | 0.4492 |
| England | Week 28 | 247361 | 0.4429 |
| England | Week 29 | 251452 | 0.4351 |
| England | Week 30 | 256074 | 0.4267 |
| England | Week 31 | 261095 | 0.4176 |
| England | Week 32 | 266919 | 0.4075 |
| England | Week 33 | 273589 | 0.3943 |
| England | Week 34 | 280330 | 0.3813 |
| England | Week 35 | 288277 | 0.3671 |
| England | Week 36 | 303489 | 0.3462 |
| England | Week 37 | 322517 | 0.3235 |
| England | Week 38 | 348347 | 0.3032 |
| England | Week 39 | 377732 | 0.2816 |

<sup>+</sup> The data of “Pillar 2” started to report from week 13th, so we started by week 13

**Fig. 1.**
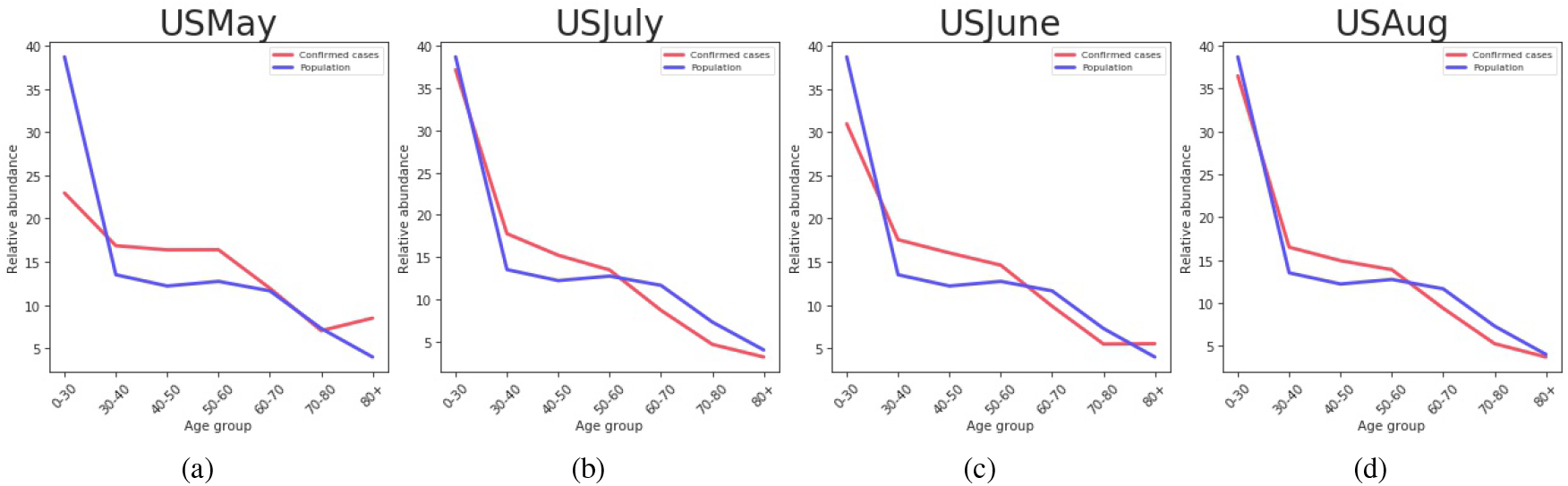
The juxtaposition of age-distribution of confirmed cases, and the US’s population for (a) May (b) June (c) July and (d) August.

**Fig. 2.**
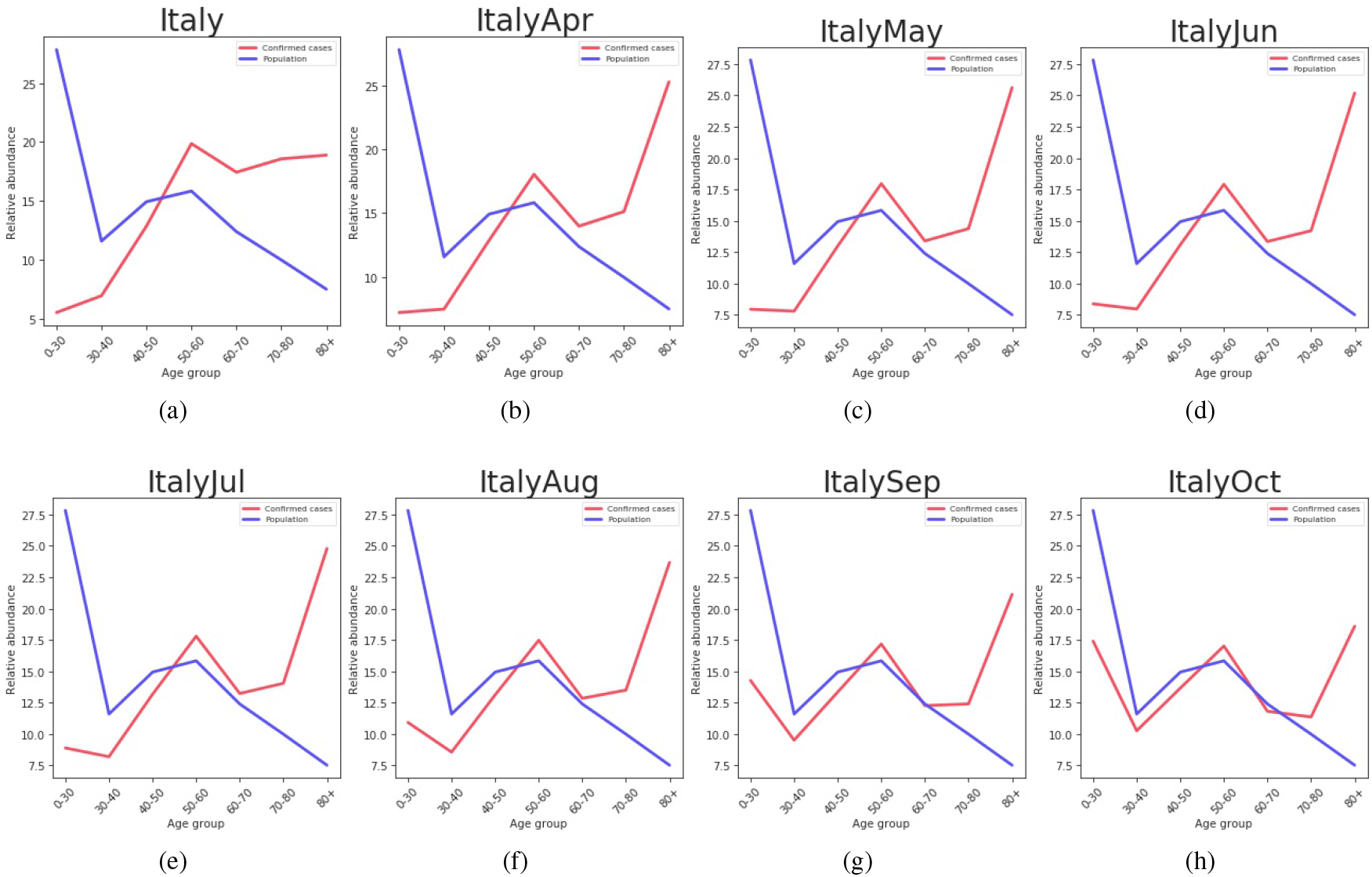
The juxtaposition of age-distribution of confirmed cases, and Italy’s population for (a) March (b) April (c) May (d) June (e) July (f) August (g) September and (h) October.

**Fig. 3.**
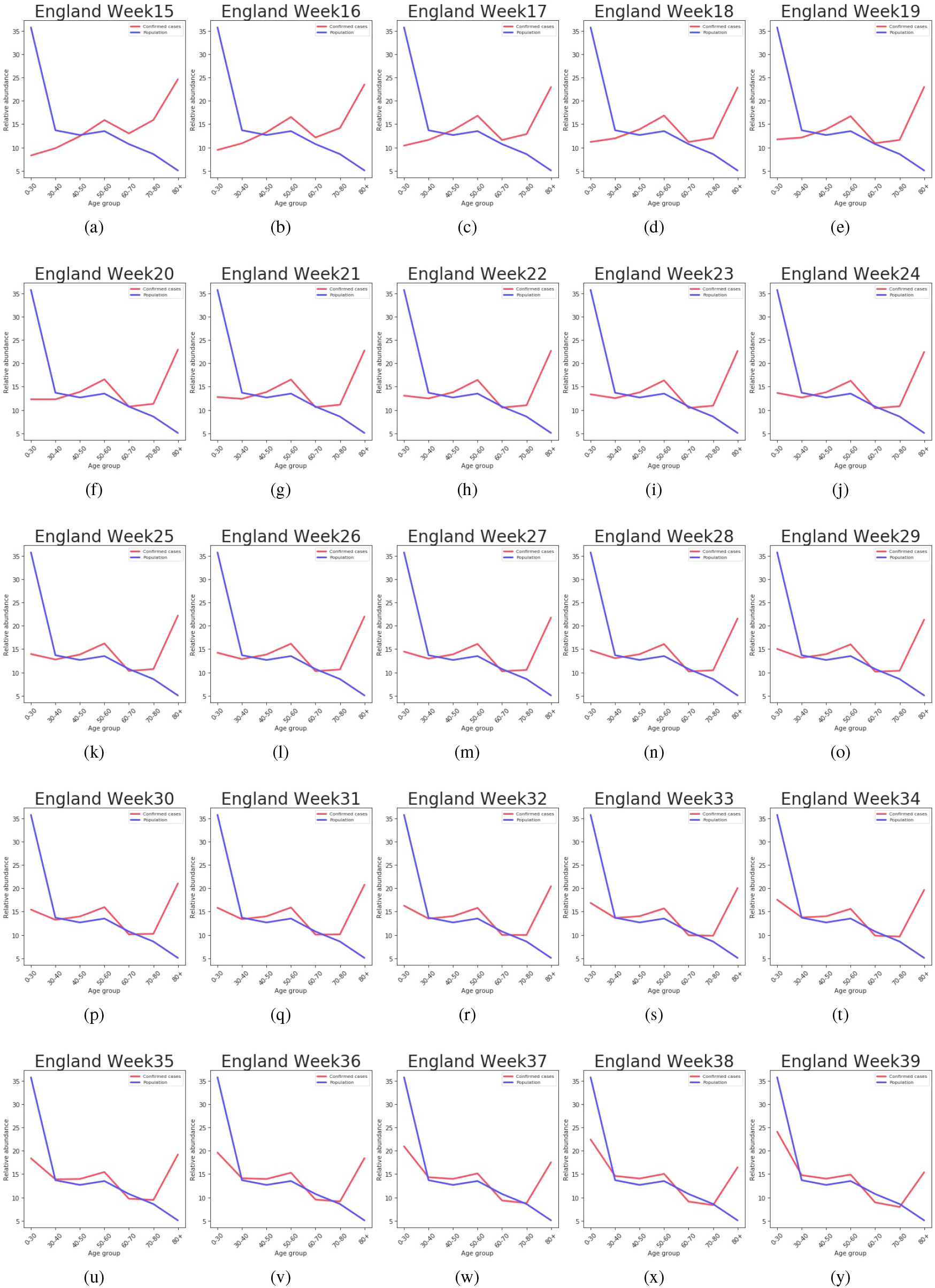
The juxtaposition of age-distribution of confirmed cases, and England’s population, for Weeks 15 to 39.

**Fig. 4.**
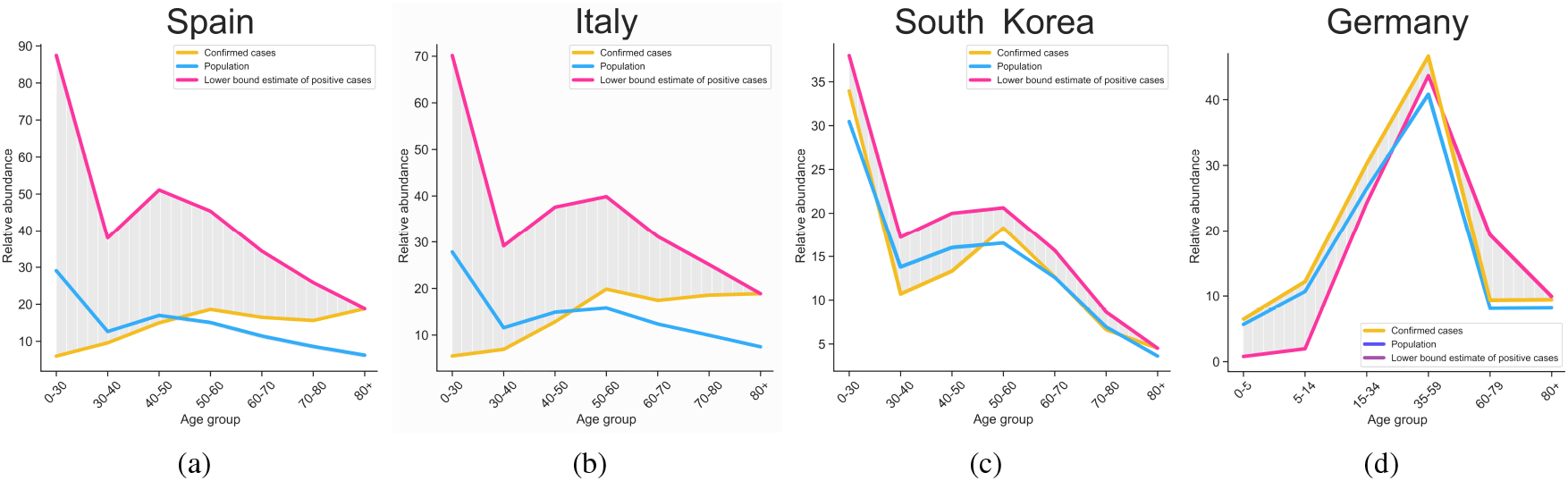
The juxtaposition of age-distribution of confirmed case, region’s population and estimated positive cases for (a) Spain (b) Italy (c) South Korea and (d) Germany.

As it expected, countries get prepared for COVID-19 and ran some strategies to both stops the spread and track more accurately the positive cases (i.e. “COVID Tracking Project” in the US), over time. Because, in the COVID-19’s cases, it is known that, positive cases with no or mild symptoms would also transmit the virus. So, testing and detecting such cases in most of the countries got more and more important over time. In addition, in many countries, in the first days of the outbreak, the number of available tests were not enough, so, it was preferred that to test people with more symptoms. As it is known today, older cases have a higher chance of severity [20], so the age-distribution of COVID-19 left-skewed in the first days of outbreak. After overcoming these problems, and over time, the skewness of the age-distribution gets reduced. This could lead us to mention that the age would not impact transmission, but the patient would be more symptomatic in older ones.

### 3.2 Lower bound estimation of positive cases

Establishing high-level health care policies (i.e. quarantine) rely on an accurate estimation of infected cases. Respiratory, contact, and aerosol transmission are the main known mechanisms by which the virus spreads. It has also been shown that the susceptibility to the infection depends mainly on viral load and exposure to an infected person [5, 7]. Therefore, and following section 3.1, the age-distribution of infected cases is expected to be similar in shape to that of the region’s population. However, we observe that, in regions with relatively higher CFR, the age-distribution of infected is skewed towards the older ages (Figure 4b,4a). Further, the observed differences between CFR of regions with similar population’s age-distributions are beyond differences in health care quality and other such contributing factors.

We hypothesize that these inconsistencies are related to how these testings are conducted and that the infection in older age groups is more symptomatic and thus more detectable. Although the asymptomatic positive cases, more prevalent among younger age groups, may go unnoticed, they equally contribute to the spread of the virus in society.

To address the aforementioned inconsistencies, the data on the oldest age group (i.e. >80-year-old group) are considered. Assuming that due to the severity of symptoms, all of the positive cases in this age group are identifiable, the ratio of positive cases over the entire population for this age group was calculated. Next, the calculated ratio was applied to the populations of other age groups to estimate a lower number for the number of positive cases for them. The reason that the obtained estimation is a lower bound of positive cases is two-fold:

1. The method assumes that all of the infected cases in the >80 age group are correctly identified, which is not necessarily the case; and
2. The members of the >80 age group, in reality, have fewer social contacts than other age groups. Thus, they have less chance of exposure to the virus than the younger population.

Lower bound estimation results are summarized in Table 4 for different regions. It is observed from the table that the ratio of the lower bound estimate of the positive case to the confirmed cases, for the two countries that have lower CFRs, namely South Korea and Germany, is very close to 1.0. This is while this ratio is much greater than 1.0, for the two countries that have higher CFRs, namely Italy and Spain. Further, Fig. 4 juxtaposes the age-distribution of the resulting lower bound estimate with that of the confirmed cases and region’s population for Italy, South Korea, Germany, and Spain. Interestingly, South Korea and Germany, which have lower CFRs, show more similarity between the age-distributions of estimated positive cases and population. On the other hand, in Italy and Spain with higher CFRs, the discrepancy between the two distributions is more strongly pronounced.

**Table 4.** The juxtaposition of the age-distribution of the resulting lower bound estimation with that of the confirmed cases and region’s population for 4 different countries

| Country | Collection Date | Confirmed Cases (CC) | Lower Bound Estimate (LBE) | LBE/CC-DCP | CFR%<sup>+</sup> | Median Age [21] |
| --- | --- | --- | --- | --- | --- | --- |
| Italy | March 31 | 94,088 | 237,086 | 2.52-0.58 | ~14.2 | 46.5 |
| Spain | April 13 | 119,994 | 360,988 | 3.01-0.56 | ~12.2 | 43.9 |
| South Korea | April 15 | 10,591 | 13,203 | 1.25-0.33 | ~2.1 | 43.2 |
| Germany | April 15 | 126,937 | 145,171 | 1.14-0.32 | ~2.6 | 47.8 |

<sup>+</sup> Calculated with regression method [22]

## 4 Conclusion

In this work, first, we compared the age-distribution of COVID-10 and the population of some countries. This leads us to find more similarities between these two distributions over time. Then, we have proposed a method for estimating a lower bound for the number of positive cases by using the reported data on the oldest age group and the regions’ population age-distributions. The proposed estimation method improved the expected similarity between the age-distribution of positive cases and the region’s population. Moreover, it was observed that regions with higher CFR show more discrepancy between the age-distribution of confirmed cases and the region’s population. This discrepancy was quantified by calculating the error of confirmed cases against our estimated lower bound.

This leads us to find a more accurate estimation of COVID-19 cases. This can help policymakers assess how the country/community is doing in regard to COVID-19 and when and how strict the mitigation policies should be. In this paper, using the publicly accessible data for several developed countries, we show how the improvement of testing over the course of several months has made it clear for the community that different age groups are equally prone to becoming COVID positive. Having knowledge of this fact helps policymakers enforce more effective policies for controlling the spread of the virus.

## Data Availability

All data are publicly available.

## References

1. Neil M Ferguson, Daniel Laydon, Gemma Nedjati-Gilani, Natsuko Imai, Kylie Ainslie, Marc Baguelin, Sangeeta Bhatia, Adhiratha Boonyasiri, Zulma Cucunubá, Gina Cuomo-Dannenburg, et al. Impact of non-pharmaceutical interventions (npis) to reduce covid-19 mortality and healthcare demand. 2020. DOI, 10:77482, 2020.

2. Matteo Chinazzi, Jessica T Davis, Marco Ajelli, Corrado Gioannini, Maria Litvinova, Stefano Merler, Ana Pastore y Piontti, Kunpeng Mu, Luca Rossi, Kaiyuan Sun, et al. The effect of travel restrictions on the spread of the 2019 novel coronavirus (covid-19) outbreak. Science, 368(6489):395–400, 2020.

3. Moritz UG Kraemer, Chia-Hung Yang, Bernardo Gutierrez, Chieh-Hsi Wu, Brennan Klein, David M Pigott, Louis Du Plessis, Nuno R Faria, Ruoran Li, William P Hanage, et al. The effect of human mobility and control measures on the covid-19 epidemic in china. Science, 368(6490):493–497, 2020.

4. Solomon Hsiang, Daniel Allen, Sébastien Annan-Phan, Kendon Bell, Ian Bolliger, Trinetta Chong, Hannah Druckenmiller, Luna Yue Huang, Andrew Hultgren, Emma Krasovich, et al. The effect of large-scale anti-contagion policies on the covid-19 pandemic. Nature, 584(7820):262–267, 2020.

5. Qun Li, Xuhua Guan, Peng Wu, Xiaoye Wang, Lei Zhou, Yeqing Tong, Ruiqi Ren, Kathy SM Leung, Eric HY Lau, Jessica Y Wong, et al. Early transmission dynamics in wuhan, china, of novel coronavirus–infected pneumonia. New England Journal of Medicine, 2020.

6. Yang Liu, Li-Meng Yan, Lagen Wan, Tian-Xin Xiang, Aiping Le, Jia-Ming Liu, Malik Peiris, Leo LM Poon, and Wei Zhang. Viral dynamics in mild and severe cases of covid-19. The Lancet Infectious Diseases, 2020.

7. Chung-Ming Chu, Leo LM Poon, Vincent CC Cheng, Kin-Sang Chan, Ivan FN Hung, Maureen ML Wong, Kwok-Hung Chan, Wah-Shing Leung, Bone SF Tang, Veronica L Chan, et al. Initial viral load and the outcomes of sars. Cmaj, 171(11):1349–1352, 2004.

8. Nicholas G Davies, Petra Klepac, Yang Liu, Kiesha Prem, Mark Jit, Rosalind M Eggo, CMMID COVID-19 working group, et al. Age-dependent effects in the transmission and control of covid-19 epidemics. medRxiv, 2020.

9. Stephen A Lauer, Kyra H Grantz, Qifang Bi, Forrest K Jones, Qulu Zheng, Hannah R Meredith, Andrew S Azman, Nicholas G Reich, and Justin Lessler. The incubation period of coronavirus disease 2019 (covid-19) from publicly reported confirmed cases: estimation and application. Annals of internal medicine, 2020.

10. Neil Ferguson, Daniel Laydon, Gemma Nedjati-Gilani, Natsuko Imai, Kylie Ainslie, Marc Baguelin, Sangeeta Bhatia, Adhiratha Boonyasiri, Zulma Cucunubá, Gina Cuomo-Dannenburg, et al. Report 9: Impact of non-pharmaceutical interventions (npis) to reduce covid19 mortality and healthcare demand. Imperial College London, 10:77482, 2020.

11. D Adam. Special report: The simulations driving the world’s response to covid-19. Nature, 2020.

12. Ensheng Dong, Hongru Du, and Lauren Gardner. An interactive web-based dashboard to track covid-19 in real time. The Lancet infectious diseases, 2020.

13. The Robert Koch Institute. Robert koch-institut: Covid-19-dashboard, 2020. Accessed: 2020/4/15.

14. Istituto Superiore di Sanita. Epidemia covid-19, 2020. Accessed: 2020/3/30.

15. Ministry of Health. Situación en españa, 2020. Accessed: 2020/4/13.

16. Korea Centers for Disease Control and Prevention. The updates on covid-19 in korea, 2020. Accessed: 2020/4/15.

17. Public Health England. National covid-19 surveillance reports, 2020. Accessed: 2020/10/20.

18. Esteban Ortiz-Ospina Max Roser, Hannah Ritchie and Joe Hasell. Coronavirus pandemic (covid-19). Our World in Data, 2020. https://ourworldindata.org/coronavirus.

19. United Nations. Population, 2020. Accessed: 2020/4/4.

20. Ryosuke Omori, Ryota Matsuyama, and Yukihiko Nakata. The age distribution of mortality from novel coronavirus disease (covid-19) suggests no large difference of susceptibility by age. Scientific reports, 10(1):1–9, 2020.

21. Centeral Intelligence Agency CIA. The world factbook, 2020. Accessed: 2020/4/15.

22. Amirhoshang Hoseinpour Dehkordi, Majid Alizadeh, Pegah Derakhshan, Peyman Babazadeh, and Arash Jahandideh. Understanding epidemic data and statistics: A case study of covid-19. arXiv preprint 2003.06933, 2020.

